# A Pragmatic Randomized Trial of an EHR-Integrated Generative AI Chart Summarization Tool for Ambulatory Clinicians

**DOI:** 10.64898/2026.08.26.26361496

**Authors:** Aaron T. Chin, Nina Zhu, Sitaram Vangala, Hawkin Woo, Lauren E. Wisk, Thomas Kingsley, John N. Mafi, Paul J. Lukac

## Abstract

**BACKGROUND:** Generative AI (genAI) chart summarization tools embedded in electronic health records (EHRs) are being rapidly deployed across U.S. health systems. Although these tools represent a promising solution to alleviate cognitive burdens, their effects have not been examined in randomized-clinical trials (RCTs).

**METHODS:** In this pragmatic RCT at a single academic health system, 284 outpatient clinicians across forty-two specialties were assigned 1:1 to Epic’s outpatient chart summarization tool or a usual-care control arm over 90 days, from February 23 to May 23, 2026. The primary outcome was physician task load (PTL) adapted for pre-charting. Prespecified exploratory outcomes included additional validated psychometrics as well as usability, safety, and time-based measures. Descriptive statistics included interaction and usage of the tool.

**RESULTS:** Of 74,474 AI chart summaries generated, 14.2% were interacted with by a clinician; the proportion of generated summaries interacted with declined from 21.5% in month 1 to 10.5% in month 3, and the proportion of clinicians using the tool at least once per month declined from 88.7% to 66.2%. The adjusted between-arm difference in PTL at follow-up favored the intervention arm (scale 0-400; −27.4; 95% CI, −49.4 to −5.3; P=0.02). Among the Professional Fulfillment Index (PFI; scale 0-4, lower=better) psychometrics, overall burnout (−0.20; 95% CI, - 0.38 to −0.01) and work exhaustion (−0.24; 95% CI, −0.47 to −0.02) were lower in the intervention arm, with little difference in overall professional fulfillment (+0.04; 95% CI, −0.16 to 0.25). Charting time per encounter showed no significant between-arm difference during steady state (−1.2 seconds; 95% CI, −19.0 to 16.6). The net promoter score was −22, indicating that on average, clinicians did not recommend the tool. Among free-text respondents, 57.1% reported at least one concern, most commonly tool limitations or inaccurate information. No adverse patient safety events or near-misses were reported.

**CONCLUSION:** An EHR-integrated AI chart summarization tool modestly reduced physician task load and was associated with lower burnout, without time savings and against declining engagement. Sustained usage and oversight of reported inaccuracies remain open challenges.

**Trial Registration:** ClinicalTrials.gov Identifier: <u>NCT07438743</u>

## Introduction

More than half of U.S. healthcare organizations have already deployed generative AI (genAI) into their electronic health records (EHR), yet rigorous evidence on whether it actually improves clinical workflows remains limited.^1,2^ These tools now support a growing range of tasks, including documentation, patient-message drafting, and discharge summaries.^3–7^ Much of this expansion is being driven by EHR vendors integrating genAI features directly into clinical workflows, with rapid market consolidation around a small number of dominant platforms.^8^ Epic Systems, the predominant U.S. EHR vendor, exemplifies this trajectory: the company disclosed that its chart-summary suite (named “Insights”) was reportedly used approximately 16 million times per month as of February 2025, roughly tripling over the preceding three months.^9^ Together, these trends suggest that EHR-integrated genAI is no longer a niche innovation or limited pilot effort; it is rapidly becoming part of everyday clinical infrastructure. EHR-integrated genAI could reduce the time required to retrieve, synthesize, and document clinical information, thereby streamlining pre-charting and improving clinician experience.

Despite this rapid and widespread implementation, the benefits of AI-based note summarization remain understudied.^6,7,10–12^ The available evidence has not kept pace with deployment, particularly for Epic-based tools that are increasingly embedded into routine workflows. GenAI outputs may include hallucinations, inaccuracies, and omissions of clinically important details, creating a potential cascade of downstream risks for clinician workload, decision-making, and patient safety.^10,13,14^ These systems also impose potentially significant financial, operational, and computational costs.^15,16^ Yet despite widespread use and high-stakes integration into care delivery, there remains little rigorous evaluation of the benefits and costs of these tools in real-world practice.

To address the lack of rigorous experimental comparisons of these novel tools against standard practices, we conducted a randomized-clinical trial (RCT) testing Epic’s ambulatory chart summarization tool (Outpatient Insights) to determine whether it meaningfully reduces physician task load (PTL) compared with usual practice.

## Methods

### Trial Design

Enrolled participants (N = 284) were randomized 1:1 to either the intervention or control group over a 90-day period from midnight of February 23 to May 23, 2026. To achieve cohort balance in terms of important baseline predictors of the primary endpoint, covariate-constrained randomization was performed. Clinicians were stratified by pre-existing access to an active AI scribe license (suggesting some degree of baseline AI literacy), and within-stratum randomization was balanced on baseline physician task load (PTL; NASA-TLX-adapted score) and baseline modified total chart time per encounter (a Caboodle-derived metric; Epic Systems, Inc., Verona, WI). A mandatory baseline pre-study survey was distributed on January 08, 2026, while the post-study survey was distributed on May 24, 2026. The intervention window was divided into three monthly periods for utilization analyses: month 1 (February 23 to March 21, 2026), month 2 (March 22 to April 18, 2026), and month 3 (April 19 to May 23, 2026). The protocol specified midnight of May 24, 2026 as the trial endpoint, which corresponds to the last full day of intervention on May 23, 2026, the date used throughout for clarity.

The study protocol, following Standard Protocol Items: Recommendations for Interventional Trials – Artificial Intelligence (SPIRIT-AI) guidelines, was published via pre-print on medRxiv and registered on ClinicalTrials.gov (NCT07438743).^17,18^ Reporting of results follows the Consolidated Standards of Reporting Trials – Artificial Intelligence (CONSORT-AI), and trial recruitment is summarized in the CONSORT-AI flow diagram (Figure 1).^19^

**Figure 1.**
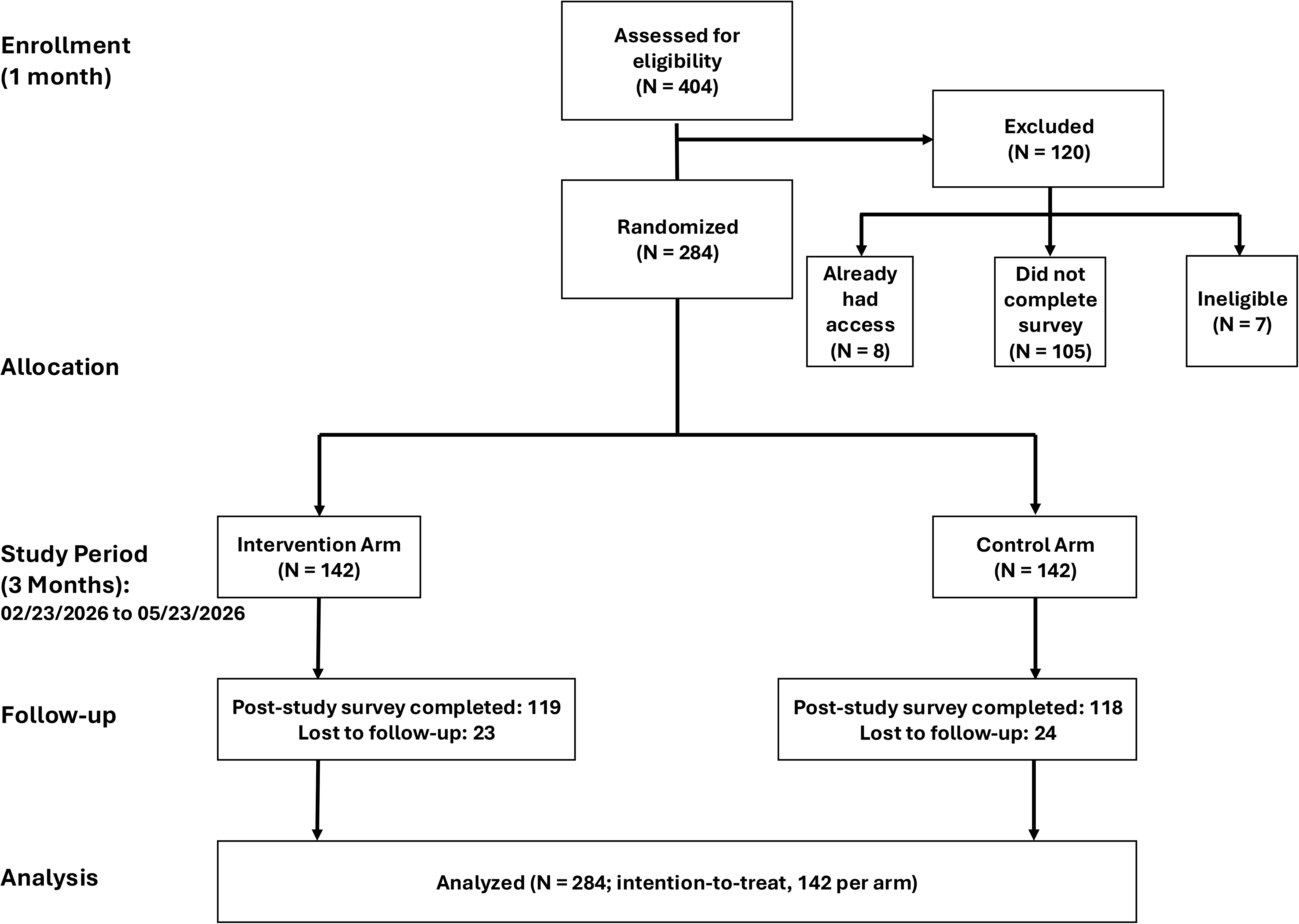
CONSORT-AI flow diagram.

### Participants

Eligible clinicians were ambulatory physicians and advanced practice providers (APPs; e.g., nurse practitioners) across forty-two specialties who held at least one half-day of clinic per week. Of 404 clinicians who expressed interest, 120 were excluded because they already had access to the tool (n=8), did not complete the baseline survey (n=105), or were ineligible (n=7) (Figure 1).

### Intervention

Intervention arm participants continued usual clinical practice with optional access to the tool, presented in an easily accessible sidebar in the patient’s chart. The tool automatically generates general or topic-focused summaries of recent clinical notes for patients on a clinician’s schedule and is intended to support chart review and visit preparation. Model details, batch and on-demand generation, input note types, input length limits, and note-selection behavior are described in the Supplement.

### Measures

We examined tool utilization at the clinician and summary levels. Summary-level counts, copy rate, and clinician-level use definitions were derived from Epic-provided reports and the Caboodle UAL.

Outcomes were drawn from survey instruments and EHR-derived metrics. Survey-based measures were collected at baseline (pre-study Qualtrics survey) and follow-up (post-study survey). EHR-derived time metrics were drawn from two Epic sources, the Caboodle User Action Log and Signal.

#### Primary outcome

The prespecified primary outcome was physician task load (PTL), adapted from the validated NASA Task Load Index and administered with items framed to the pre-charting context (0-400 scale, where lower scores indicate less cognitive load).^20^ We selected PTL as the primary outcome because our conceptual model of the tool’s mechanism of action centers on reducing cognitive burden associated with chart review, potentially saving the clinician mental capacity for other activities.^20^

#### Pre-specified exploratory outcomes

Prespecified exploratory outcomes included additional survey measures as well as EHR time-based efficiency metrics.

The Professional Fulfilment Index (PFI) assesses professional fulfillment and overall burnout (0-4 scale, where higher scores for professional fulfillment indicate higher professional fulfilment and higher burnout scores indicate higher burnout).^21^ Three additional post-study novel survey items, each rated on a five-point Likert scale, assessed clinicians’ perceptions of their pre-charting effectiveness.

EHR-derived time was measured with two metrics. Modified total chart time per encounter, derived from the User Action Log (UAL) within Caboodle (Epic’s enterprise data warehouse), quantified the time clinicians spent reviewing patient charts prior to the end of an encounter. This custom metric was internally validated because Epic’s Chart Review Signal metric did not adequately capture chart review time for the purposes of this study (Supplement). The Epic Signal metric “time outside scheduled hours” was captured to account for pre-charting that occurs outside of a clinician’s clinical hours. Baseline values for both were drawn from the six months preceding the intervention.

Usability and user satisfaction were measured using the system usability scale (SUS) and the net promoter score (NPS). The SUS’s 10-item questionnaire evaluated how comfortable clinicians were using the tool (0-100 scale, score less than 50 suggests poor usability).^22^ The NPS summarizes consumer satisfaction using a single question asking respondents how likely they are to recommend the tool, classifying them as detractors (0-6), passives (7-8), or promoters (9-10), with NPS calculated as percentage of promoters minus percentage of detractors (scores −100 to 100, higher score indicates higher satisfaction).^23^

The post-study survey also assessed prespecified safety-related topics using five-point Likert items, including clinician-perceived frequency of clinically significant inaccuracies and biases and the occurrence of adverse events or near misses.^24^ Free-text feedback was obtained from two sources: an open-response post-study survey item asking clinicians to describe any tool-generated inaccuracies or biases they observed, and unsolicited feedback submitted through the tool’s embedded feedback mechanism during the intervention period. Free-text responses were categorized into mutually exclusive content categories using an inductively developed coding scheme (Supplement).

### Statistical Methods

Clinician-period level linear mixed models were used to estimate intervention effects, with random clinician effects accounting for repeated measurements. Models included terms for arm, period, and their interaction, and adjusted for prespecified baseline characteristics (age, sex, specialty, number of clinic half-days per week, baseline PTL, baseline modified total chart time per encounter, and ambient scribe access). For survey outcomes, the included periods were baseline and follow-up, while for the EHR metrics, these were baseline, month 1 of follow-up (learning curve), and month 2 and 3 combined (steady state). Linear contrasts at follow-up (survey) or steady state (EHR) were used for intervention effect estimation.

Under the missing-at-random assumption, the prespecified intention-to-treat (ITT) analysis of survey outcomes was robust to loss to follow-up. The primary outcome was evaluated at a two-sided significance level of 0.05. Alpha was entirely reserved for the primary outcome; hence no method for adjustment of multiplicity across the prespecified exploratory outcomes was specified in the protocol or statistical analysis plan. Consistent with this, all exploratory outcomes are reported as effect estimates with 95% confidence intervals, without P values. The widths of these confidence intervals have not been adjusted for multiplicity, and the inferences drawn from them may not be reproducible. Analyses were performed using R v. 4.5.1. Full model specifications, sample size justification, and adjustment covariates are detailed in the Supplement.

## Results

### Participants

A total of 284 clinicians were enrolled (Table 1). Approximately half of clinicians in both arms were 35 to 44 years old (46.5% intervention, 50.7% control), and 83.1% of the intervention arm and 83.2% of the control arm held an active ambient AI scribe license. The post-study survey was completed by 119 of 142 (83.8%) intervention-arm clinicians and 118 of 142 (83.1%) control-arm clinicians. Of the 119 intervention-arm clinicians who returned the post-intervention survey, 118 answered the tool-specific frequency items and NPS item, and 116 completed all 10 System Usability Scale items.

**Table 1.**
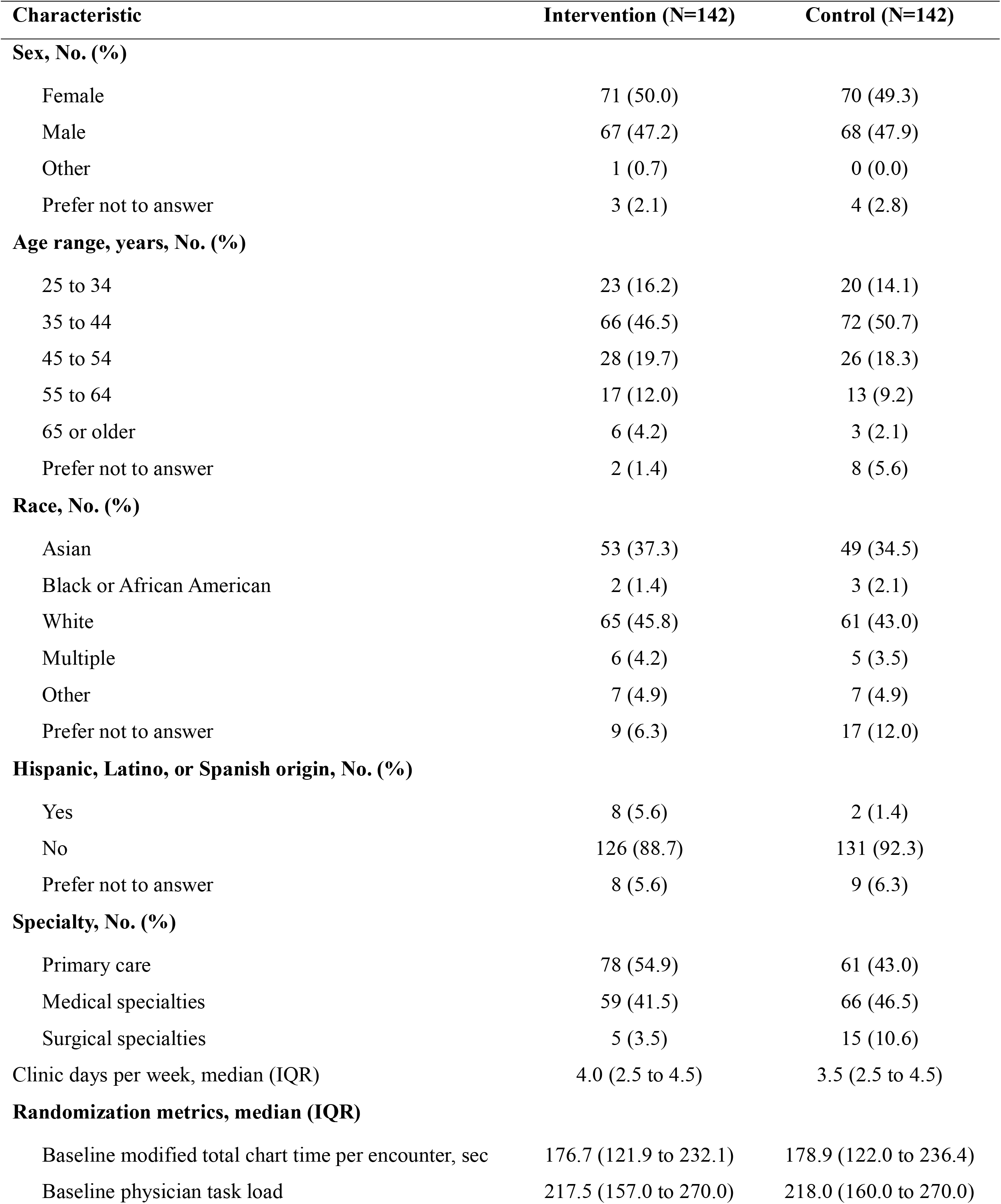
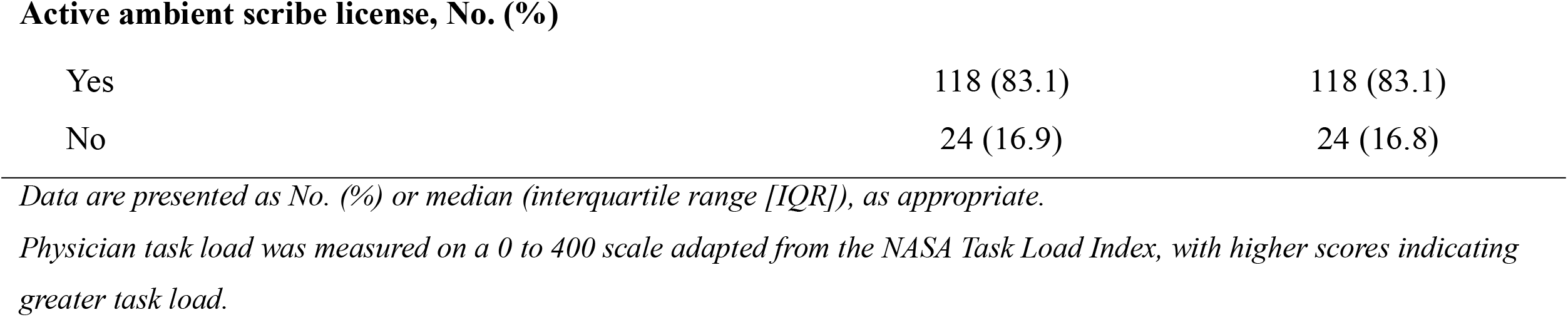
Baseline characteristics of participants by study group.

| Characteristic | Intervention (N=142) | Control (N=142) |
| --- | --- | --- |
| **Sex, No. (%)** |  |  |
| Female | 71 (50.0) | 70 (49.3) |
| Male | 67 (47.2) | 68 (47.9) |
| Other | 1 (0.7) | 0 (0.0) |
| Prefer not to answer | 3 (2.1) | 4 (2.8) |
| **Age range, years, No. (%)** |  |  |
| 25 to 34 | 23 (16.2) | 20 (14.1) |
| 35 to 44 | 66 (46.5) | 72 (50.7) |
| 45 to 54 | 28 (19.7) | 26 (18.3) |
| 55 to 64 | 17 (12.0) | 13 (9.2) |
| 65 or older | 6 (4.2) | 3 (2.1) |
| Prefer not to answer | 2 (1.4) | 8 (5.6) |
| **Race, No. (%)** |  |  |
| Asian | 53 (37.3) | 49 (34.5) |
| Black or African American | 2 (1.4) | 3 (2.1) |
| White | 65 (45.8) | 61 (43.0) |
| Multiple | 6 (4.2) | 5 (3.5) |
| Other | 7 (4.9) | 7 (4.9) |
| Prefer not to answer | 9 (6.3) | 17 (12.0) |
| **Hispanic, Latino, or Spanish origin, No. (%)** |  |  |
| Yes | 8 (5.6) | 2 (1.4) |
| No | 126 (88.7) | 131 (92.3) |
| Prefer not to answer | 8 (5.6) | 9 (6.3) |
| **Specialty, No. (%)** |  |  |
| Primary care | 78 (54.9) | 61 (43.0) |
| Medical specialties | 59 (41.5) | 66 (46.5) |
| Surgical specialties | 5 (3.5) | 15 (10.6) |
| Clinic days per week, median (IQR) | 4.0 (2.5 to 4.5) | 3.5 (2.5 to 4.5) |
| **Randomization metrics, median (IQR)** |  |  |
| Baseline modified total chart time per encounter, sec | 176.7 (121.9 to 232.1) | 178.9 (122.0 to 236.4) |
| Baseline physician task load | 217.5 (157.0 to 270.0) | 218.0 (160.0 to 270.0) |
| **Active ambient scribe license, No. (%)** |  |  |
| Yes | 118 (83.1) | 118 (83.1) |
| No | 24 (16.9) | 24 (16.8) |

- *Data are presented as No. (%) or median (interquartile range [IQR]), as appropriate.* *Physician task load was measured on a 0 to 400 scale adapted from the NASA Task Load Index, with higher scores indicating* *greater task load.*

### Usage

A total of 74,474 summaries were generated across the trial, of which 10,591 (14.2%) were interacted with by a clinician (defined as manual generation or an auto-generated summary with recorded clinician interaction; Figure 2A). Interacted-with summaries comprised 7,120 manually generated summaries (9.6% of generated) and 3,471 auto-generated summaries with recorded clinician interaction (4.7% of generated). The proportion of generated summaries interacted with declined from 21.5% in month 1 to 12.0% in month 2 and 10.5% in month 3. The proportion of intervention-arm clinicians who used the tool at least once during each month declined from 88.7% (95% CI, 82.5 to 92.9) in month 1 to 77.5% (95% CI, 69.9 to 83.6) in month 2 and 66.2% (95% CI, 58.1 to 73.5) in month 3 (Figure 2B). Of the 10,591 interacted-with summaries, 2,376 (22.4%) had content copied into the clinical note. The copy rate among interacted-with summaries rose across months, from 17.7% in month 1 to 24.9% in month 2 and 27.3% in month 3 (Figure 2C).

**Figure 2.**
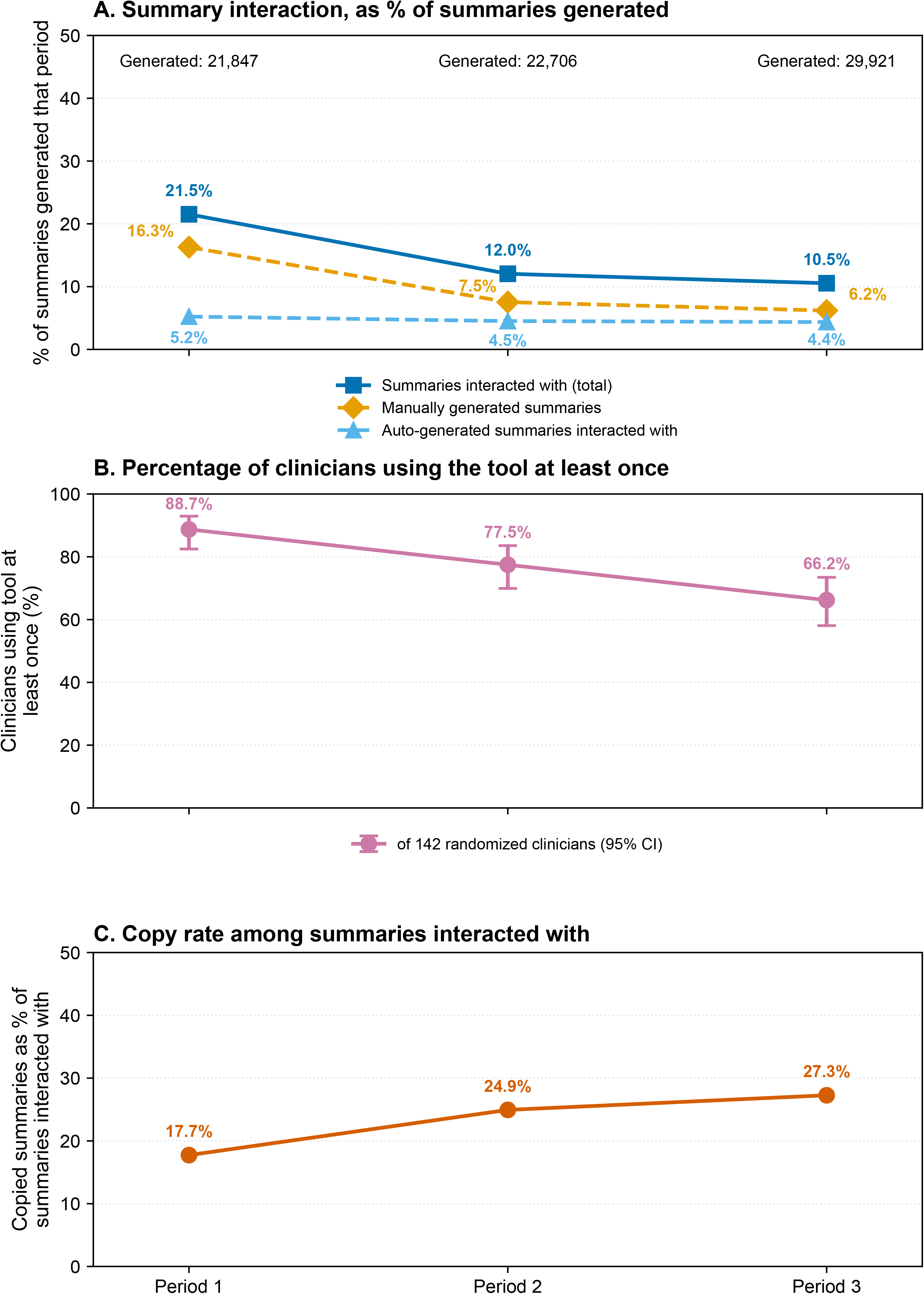
Tool utilization across the three study months. Panel A shows summary interaction volume as a percentage of total summaries generated in each month, broken into total interacted with (dark blue), manually generated (gold), and auto-generated with recorded clinician interaction (light blue). The total number of summaries generated in each month is shown above the plot. Panel B shows the percentage of intervention-arm clinicians (n=142) who used the tool at least once during each month, defined as any recorded interaction with the tool in the Caboodle User Action Log. Error bars indicate Wilson 95% confidence intervals. Panel C shows the percentage of summaries interacted with in each month that had any content copied into the clinical note.

### Primary Outcome

Mean PTL decreased from 210.1 (SD, 87.5) at baseline to 193.4 (SD, 91.7) at follow-up in the intervention arm and increased from 210.9 (SD, 89.7) to 218.2 (SD, 86.7) in the control arm. The adjusted between-arm difference at follow-up was −27.4 points (95% CI, −49.4 to −5.3; P=0.02) (Table 2).

**Table 2.**
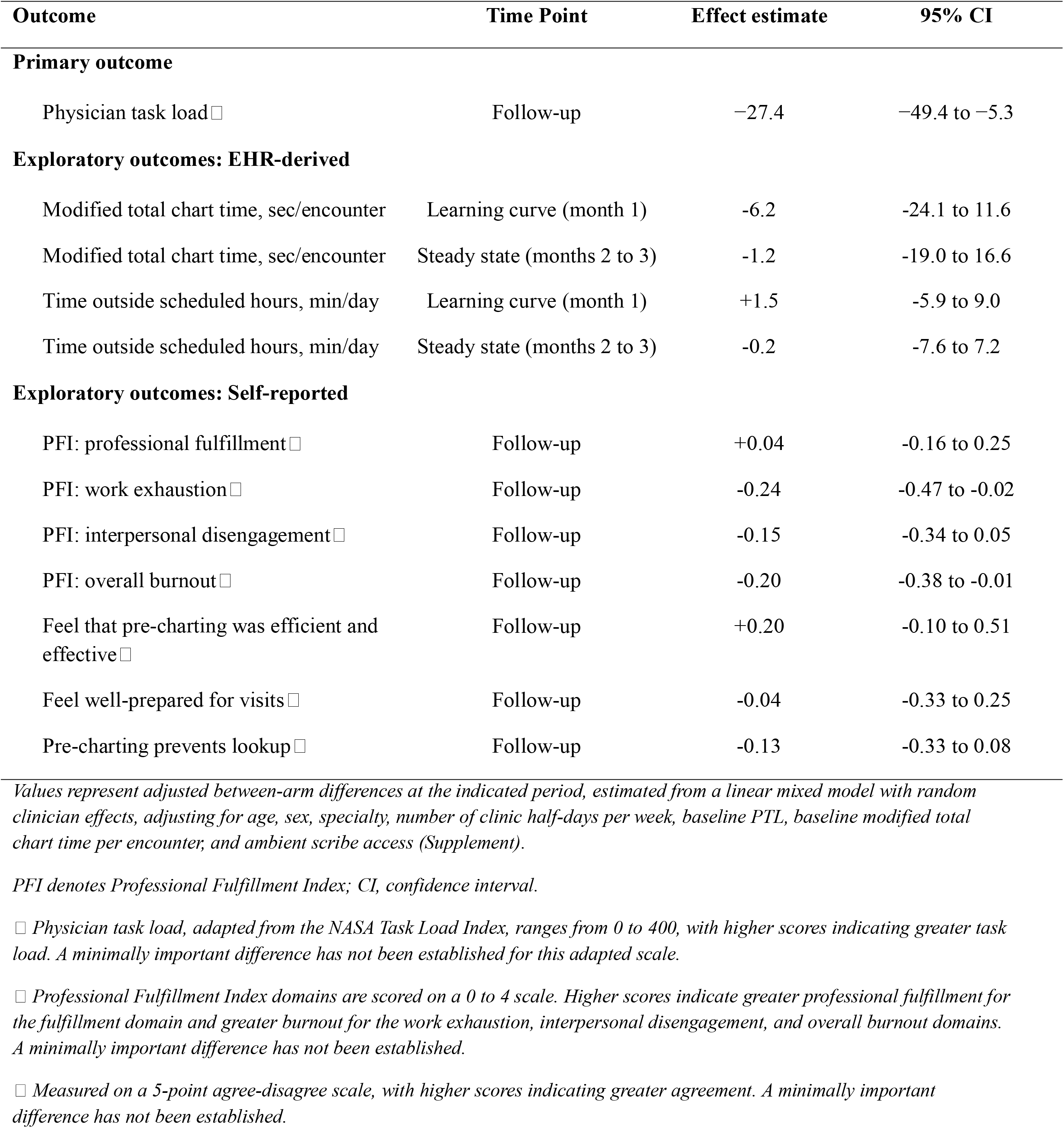
Adjusted effects of the chart summarization tool on physician task load, pre-charting time, and professional fulfillment.

### Prespecified Exploratory Outcomes

#### Professional Fulfillment Index

Overall professional fulfillment showed little between-arm difference at follow-up (adjusted difference, +0.04; 95% CI, −0.16 to 0.25). Overall burnout was lower in the intervention arm (−0.20; 95% CI, −0.38 to −0.01), as was the work exhaustion subscale (−0.24; 95% CI, −0.47 to - 0.02). The between-arm difference for interpersonal disengagement was the same directionally, but the CI included zero (−0.15; 95% CI, −0.34 to 0.05). The proportion of clinicians meeting the criterion for burnout (overall burnout ≥1.33) rose from 41.5% to 55.6% in the control arm and from 42.3% to 44.9% in the intervention arm.

#### Pre-Charting Perceptions

Between-arm differences on the three pre-charting perception items were small and crossed the null, with adjusted differences of −0.04 for feeling well-prepared for visits (95% CI, −0.33 to 0.25), −0.13 for agreement that pre-charting prevents lookup during visits (95% CI, −0.33 to 0.08), and +0.20 for agreement that pre-charting was efficient and effective (95% CI, −0.10 to 0.51).

#### Time Outcomes

Modified total chart time per encounter showed an adjusted between-arm difference of −6.2 seconds during month 1 (95% CI, −24.1 to 11.6) and −1.2 seconds during steady state (95% CI, - 19.0 to 16.6). Time outside scheduled hours showed adjusted between-arm differences of +1.5 minutes per day during month 1 (95% CI, −5.9 to 9.0) and −0.2 minutes per day during steady state (95% CI, −7.6 to 7.2).

#### Usability and Net Promoter

Item-level distributions for the SUS, tool-specific frequency items, and NPS are shown in Figure 3. The SUS produced a mean total score of 74.1 (SD, 16.3; n=116). For NPS, 118 clinicians responded: 57 (48.3%) were classified as detractors (score 0 to 6), 30 (25.4%) as passives (7 to 8), and 31 (26.3%) as promoters (9 to 10), yielding an NPS of −22.

**Figure 3.**
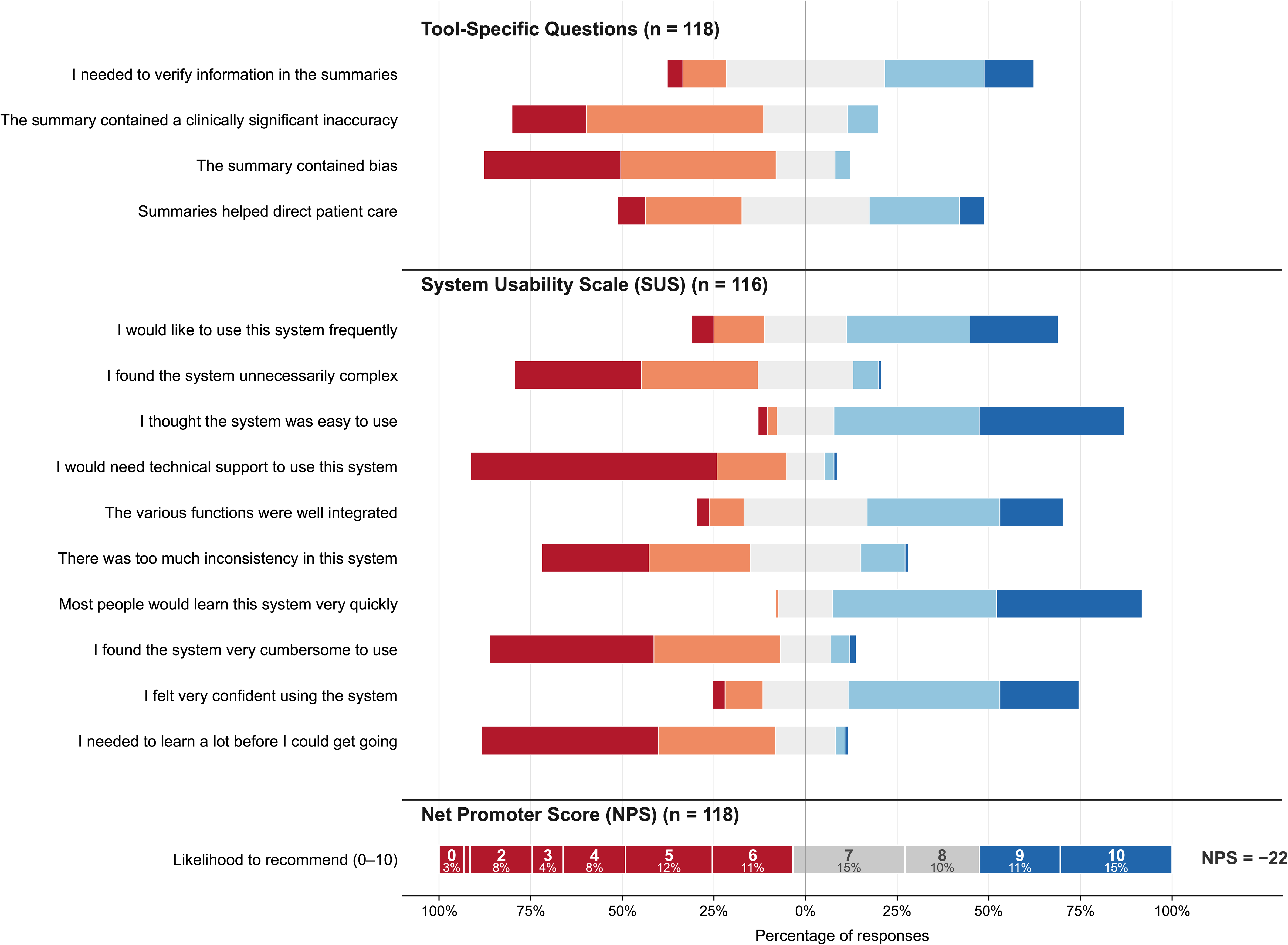
Post-intervention survey responses. Diverging stacked bars of intervention-arm clinician responses to tool-specific frequency items (Never to Always), the 10 System Usability Scale (SUS) agreement items (Strongly disagree to Strongly agree), and the Net Promoter Score (NPS) item, in a red-to-blue palette centered on the neutral category. For the NPS item, bars are colored by category (detractors 0 to 6, passives 7 to 8, promoters 9 to 10) and labeled with the percentage of respondents at each 0 to 10 score.

When asked to identify the contexts in which the summarization tool was most useful (up to two selections permitted), clinicians most frequently selected medically complex patients (63 selections) and visits with new patients (61), with fewer selections for visits with established patients (28), less medically complex patients (25), and socially complex patients (11) (Figure S1).

#### Safety and Qualitative Sentiment

Bias was reported as never or rarely occurring by 79.7% of 118 clinicians (median, 2 [IQR, 1 to 2]; scale, 1=never to 5=always), and clinically significant inaccuracy by 68.6% (median, 2 [IQR, 2 to 3]). On the corresponding verification frequency item (n=118), 19 clinicians (16.1%) reported needing to verify summary contents never or rarely, 51 (43.2%) reported sometimes, and 48 (40.7%) reported often or always (median, 3 [IQR, 3 to 4]).

In the post-intervention survey free-text item, 52 of 91 interpretable responses (57.1%) reported at least one concern (Table 3). The most frequently coded category was tool limitations (18 of 52), most often referring to the inability to summarize discrete data such as laboratory or imaging results, followed by inaccurate information (16 of 52). Reported inaccuracies included attribution errors, in which a diagnosis documented for a family member was represented as belonging to the patient, and status errors, in which an ordered but incomplete action was represented as completed.

**Table 3.** Qualitative Categories of Concerns Reported in the Post-Intervention Survey.

| Category | No. of clinicians<br>(N=52) | Representative comment |
| --- | --- | --- |
| Tool limitations | 18 | “The lack of inclusion of media and imaging for my line of work in obstetrics and maternal–fetal medicine leads to inaccuracies related to pregnancy encounters.” |
| Inaccurate information | 16 | “The HCC billing code is being misinterpreted as hepatocellular carcinoma. At times, patients were also mislabeled as having cholangiocarcinoma.” |
| Generality and relevance | 12 | “It was not necessarily inaccuracies; rather, the summary emphasized elements of the history or status that were not relevant.” |
| Trust barriers | 3 | “I am concerned that if I do not double-check the information, I may accidentally incorporate inaccurate information.” |
| Omissions | 2 | “Missed a surgery.” |
| Workflow and speed | 1 | “I kind of gave up on it after I realized it was not saving me that much time.” |

Free-text responses were collected from intervention-arm clinicians using the survey item, “Please share any examples of tool-generated inaccuracies or biases you observed, including whether specific issues occurred consistently.” Of 95 clinicians who responded, 4 responses were uninterpretable and were excluded. Among the 91 responses, 35 (38.5%) reported no observed inaccuracies or biases, 4 (4.4%) comment not addressing accuracy or bias, and 52 (57.1%) reported at least one concern. Each response was assigned to a single, mutually exclusive category. Quotations were lightly edited for spelling, punctuation, and clarity without changing meaning.

In-tool feedback submissions entered by clinicians at the time of encounter (n=72) most commonly cited inaccurate information (30 of 72 [41.7%]) and tool limitations (20 of 72 [27.8%]; Table S1). Omissions were more prominent in this unsolicited feedback (15 of 72 [20.8%]) than in the post-intervention survey (2 of 52). Flagged omissions included clinically significant recent events such as new high-impact diagnoses (for example, new cancer diagnoses or new hilar masses) and recent hospital admissions. No adverse patient safety events or near-misses were reported.

## Discussion

In this randomized-clinical trial of an outpatient EHR-integrated generative AI chart summarization tool, clinician task load significantly decreased relative to control. Despite task load benefits, usage decreased over the course of the three-month trial and users reported a negative net promoter score. These findings suggest a decoupling of measured cognitive benefit from sustained use and promotion of the tool. While no major patient safety events or near misses were noted, reported inaccuracies will necessitate ongoing clinician vigilance, particularly as genAI tools are increasingly implemented across a growing number of U.S. health systems.^1^ Lastly, the divergence between the volume of summaries generated and actual clinician engagement highlights a critical yet easily overlooked cost driver that health systems must account for as they scale adoption of the many emerging genAI features billed per -token or - generation.

The benefits reported here reflect an emerging pattern across early genAI tools in clinical workflows, in which reductions in task load, work exhaustion, and burnout coexist with minimal or no measurable time savings.^4,25–27^ An observational study of Epic’s outpatient chart summarizer demonstrated a consistent pattern, with no aggregate time savings but modest reductions among clinicians with longer baseline pre-charting times.^7^ The improvement in work exhaustion and overall burnout is consistent with prior work identifying task load as a mediator between EHR usability and burnout, in which each 10% decrease in task load was associated with lower odds of burnout in a national physician sample.^20^ In the present trial, clinicians reported the greatest perceived benefit for medically complex and new patients; encounters in which chart review demands are highest. Prior work has shown that adapted LLMs can match or exceed medical experts in clinical text summarization, providing a technical basis for benefit even when total charting time is unchanged.^28^ One explanation for the discordance between task load and time savings is that reduced cognitive burden is redirected toward other cognitive activity, such as deeper clinical reasoning or reviewing other clinical data sources, rather than faster completion.^29^ In addition, having access to a summary may enhance clinicians’ sense of pre-visit preparedness that has been associated with higher clinician satisfaction and lower burnout.^30^ Neither of these hypotheses were directly testable in this trial and thus warrant direct evaluation in future work.

The divergence between measurable task load benefit and low adoption, together with a negative NPS, may reflect a gap between the tool’s actual performance and clinicians’ expectations of it. On structured survey items, few clinicians reported that the summaries were inaccurate or biased, and most indicated they would continue to use it frequently. Aggregate ratings, however, may not fully capture how clinicians form trust in AI tools. Because trust can be shaped by granular experience with tool outputs, singular experiences with errors can disproportionately erode willingness to rely on the tool relative to its overall performance.^31^ Clinicians most often cited general limitations of the tool and inaccurate information as concerns. Although omissions were infrequent on the post-intervention survey, unsolicited in-tool feedback captured at the point of care surfaced clinically significant examples, including a summary that omitted a major new cancer diagnosis. Even rare errors of this severity may raise the perceived need to verify tool output. In this trial, a substantial proportion of clinicians reported frequent verification of summary contents. Complaints that summaries were too general or vague may also have prompted additional re-prompting, a possible contributor to workflow burden. Taken together, positive perceptions of the tool and a measurable reduction in PTL did not translate into endorsement, perhaps reflecting a threshold effect in which infrequent but salient errors outweighed the tool’s aggregate benefits.

Summarization quality is further affected by the quality of the source notes. Note bloat and copy-forward content in modern EHRs limits achievable performance regardless of model capability.^32^ Without access to structured data, the tool lacks context awareness advantageous for focused ambulatory visit. Newer summarization versions will incorporate discrete EHR data, such as lab results, but whether they can adequately contextualize these data within a narrative summary remains an open question.

These effects may also help explain the declining pattern of adoption. Utilization decreased over the three-month trial, consistent with prior evidence of modest real-world uptake of generative AI tools within the EHR.^5,26^ The declining usage over the three-month trial period contrasts with a prior deployment of a generative AI message drafting tool in which utilization increased over time following iterative prompt refinement.^5^ Tools that require separate workflow steps either provide less immediately apparent value, or lack iterative refinement based on user feedback, and ultimately may be more susceptible to declining utilization.^33^ This tool, in which the automatically generated summaries where prompts are controlled by Epic, did not undergo localized refinement, which may have contributed to its decreasing use over time. This may have been particularly relevant if clinicians perceived the tool’s outputs as insufficiently accurate or useful to justify the effort required to engage with it.^13^

A widely raised concern is that LLMs may inflate operational costs in healthcare, particularly under per-token pricing models that represent an emerging expenditure paradigm for health systems.^15,34^ In this trial, the tool was configured per Epic’s recommendation to automatically generate summaries as a batch job for all scheduled visits. With declining tool utilization, batch generation of summaries for every encounter becomes progressively harder to justify, since a growing share of generated summaries is never used. End-user manual summary generation or automatic batch generation in a subset of specialties or visit types represent alternative, cost-conscious approaches. Manual, on-demand generation would reduce cost but likely at the expense of usability, since the tool’s value depends in part on summaries being immediately available at the point of care. Given limited reimbursement pathways for most clinical AI applications, costs must be weighed against sustained utilization and demonstrated clinical value.^10^ The optimal configuration to maximize usage of this tool, and many others like it, requires ongoing study.

### Strength and Limitations

This trial has several strengths. Randomization with covariate constraint on baseline task load and chart review time minimized selection bias and confounding, which is particularly relevant for an optional workflow tool.^4^ The combination of validated instruments, EHR-derived time metrics, and structured and free-text feedback permitted evaluation across a broad array of subjective and objective dimensions. Finally, the trial evaluated a vendor-native tool already widely deployed, responding to calls for randomized evaluation of EHR-embedded generative AI applications.^10^

This trial also has limitations. First, the 90-day intervention period may not capture the full adoption curve or stable long-term workflows, and effects during early adoption may overestimate or underestimate steady-state impact. A short duration is also partially protective, as EHR-integrated generative AI tools evolve rapidly and longer trials risk evaluating a superseded version of the technology.^14^ Second, alternative configurations of the tool were not piloted, including a bulleted summary format that was available during the trial period. Many AI tools will ultimately offer a degree of user-based customizability that trials should account for. Third, summary-level interaction was measured using Epic-provided metrics that combined recorded interactions with auto-generated summaries and counts of manually generated summaries. Auto-generated summaries were counted as interacted with only when the clinician actively clicked into them, which may under-capture usage, as clinicians who viewed a summary in the sidebar without clicking would not be counted. Still, the temporal decline across periods shows a quantifiable decrease in usage. Fourth, the trial was conducted at a single academic health system using one vendor’s tool, and findings may not generalize to other settings, EHR platforms, or summarization products. Our cohort was approximately half female (49.6%), compared with 38.7% in the national ambulatory physician workforce, which may further limit generalizability.^35^ Fifth, survey-based outcomes are subject to nonresponse bias despite high completion rates, and the missing at random assumption underlying the ITT analysis cannot be verified directly.

Future research should incorporate qualitative and behavioral science methods to characterize how clinicians engage with generative AI-generated summaries during chart review, given that measurable psychometric benefit was observed without a corresponding reduction in chart review time. This pattern suggests that reduced cognitive burden may be redirected toward other cognitive activity rather than eliminated, a hypothesis that structured time-motion metrics alone cannot capture. These tools will also change as newer GPT models are introduced into the system, requiring ongoing monitoring. Larger, multicenter randomized trials are also needed to evaluate the effect of generative AI chart summarization on downstream outcomes including quality of care, cost, and health equity, outcomes that a single-center, rapid-cycle trial focused on clinician-reported measures was not designed to capture.

## Conclusion

In this randomized-clinical trial evaluating EHR-embedded, genAI-mediated chart summarization tool, we observed modest psychometric benefits without improvement in time savings. The negative NPS, alongside declining usage, suggests that the tool’s value likely lies in select use cases rather than as a universal solution. Further study is needed to identify the scenarios in which the tool performs best so that future implementation can target the highest yield applications.

## Acknowledgements

Yan Phipps for data curation. Artem Romanov for project management support. Drs. Kenny Leung and Thalia Nguyen for the modified Caboodle validation. Dominic Parsia for Epic support.

## Declarations

### Ethics approval and consent

This study was approved by the University of California, Los Angeles Institutional Review Board (IRB-26-0066). Participating clinicians provided informed consent by completing the baseline survey, which included an information sheet describing study procedures, risks, participant rights, and the voluntary nature of participation. The trial was conducted in accordance with the Declaration of Helsinki.

### Trial registration

This trial was registered at ClinicalTrials.gov on February 22, 2026 (NCT07438743). The full trial protocol is available on medRxiv at https://www.medrxiv.org/content/10.64898/2026.02.20.26346503v2. The initial protocol was posted on February 22, 2026, prior to trial initiation. A revised protocol was posted on May 24, 2026, coinciding with the scheduled end of the intervention period and prior to any analysis of trial data.

### Data availability

Deidentified aggregate data supporting the findings of this study are available from the corresponding author on reasonable request. Individual-level clinician survey responses and EHR-derived utilization data cannot be shared publicly because of institutional data governance policies and to protect participant confidentiality, given the small clinician cohort and multiple specialties.

### AI-use disclosure

The authors used a large language model (Claude, Anthropic) to assist with editorial revisions, structural feedback, and reference formatting checks. The authors reviewed and take responsibility for all content.

### Author approval

All authors reviewed and approved the final version of the manuscript prior to submission.

---

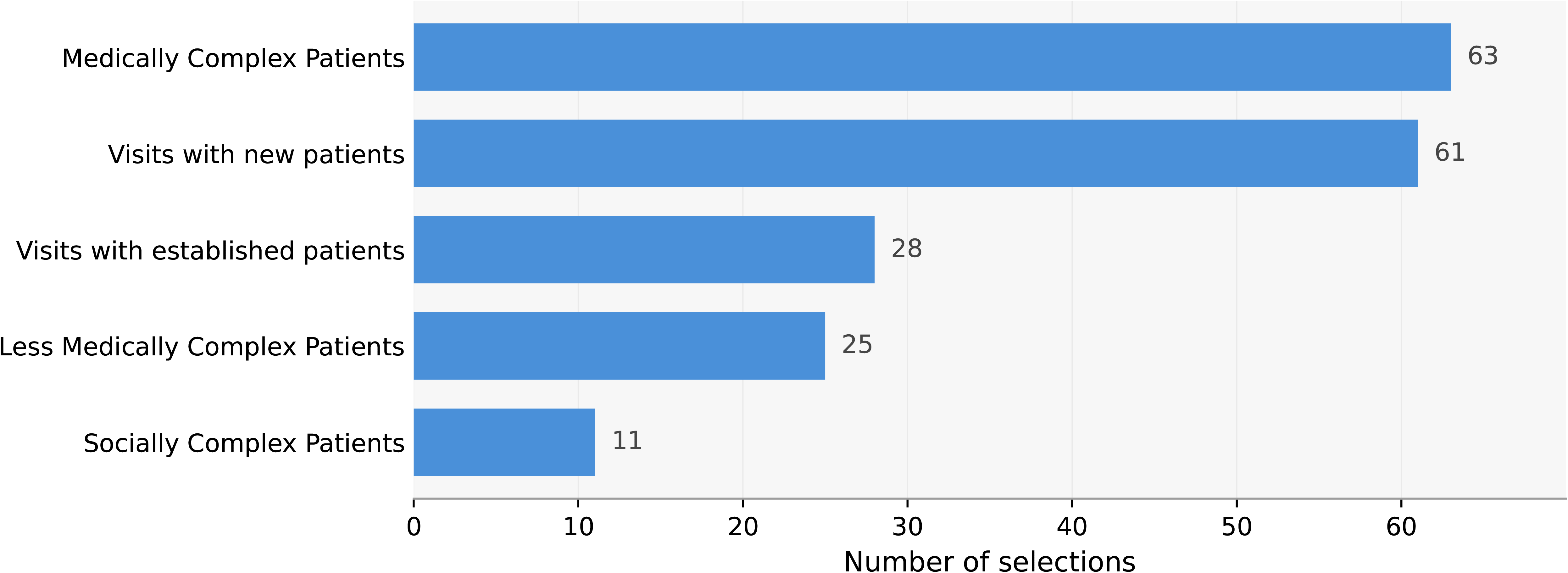

