## Supplemental for "A Pragmatic Randomized Trial of an EHR-Integrated Generative AI Chart Summarization Tool for Ambulatory Clinicians"

**Supplementary Appendix**

**Table of Contents**

| **Section** | **Page** |
| --- | --- |
| **Methods Section S1. Participants** | **2** |
| **Methods Section S2. Intervention** | **2** |
| **Methods Section S3. Measures** | **3** |
| **Methods Section S4. Statistical Analysis** | **4** |
| **Table S1. Categorization of In-Tool Feedback Submissions from Intervention-Arm Clinicians** | **5** |
| **Figure S1. Contexts in which the summarization tool was reported most useful** | **6** |

**Methods**

**Section S1. Participants**

Clinicians were recruited through department-wide emails distributed by department leaders. The 42 specialties were grouped into three categories for adjustment and stratified analysis: primary care, medical specialty, and surgical specialty.

**Section S2. Intervention**

S2.1 Participant support. All participants watched a 2-minute training video before accessing the tool and were discouraged from copying generated summaries into clinical notes. Intervention-arm participants additionally received an emailed tip sheet and were invited to four weekly virtual town halls during the first month.

S2.2 The Outpatient Insights tool used a combination of GPT-4.1-nano and GPT-4.1 to complete distinct, sequential functions to create the narrative summary. The model versions and architecture remained unchanged for the duration of the study.

S2.3 Summary generation. Summaries were generated automatically in batches approximately 36 hours before scheduled visits. Users initiated generation manually for same-day visits. A "focus on" prompt allowed users to request a topic-oriented summary from a short free-text description (e.g., "focus on the patient's asthma history"). Batch and manual generation summarized the most recent eligible notes chronologically; on-demand users could self-select notes.

S2.4 Input constraints. Summaries were derived exclusively from clinical notes and did not incorporate other EHR data such as laboratory or imaging results. Input was limited to approximately 24,000 English characters or 30 notes, whichever came first.

**Section S3. Measures**

S3.1 Tool utilization metrics. Summary-level counts were extracted from three Epic-provided reports covering the 90-day intervention window. The first captured auto-generated summaries with recorded clinician interaction, based on Epic's internal definition of interaction with a generated summary. The second captured manually generated summaries linked to a clinical encounter. The third captured manually generated summaries without a linked encounter (for example, chart review viewing outside a scheduled visit). The sum of these three streams comprised the total 'summaries interacted with' metric; manually generated summaries were counted as interacted-with by construction. Copies were detected by Epic using a proprietary summary-level mechanism; the specific triggering event was not accessible to the study team. The copy rate reported in the main text is the number of summaries with any content copied into the clinical note divided by summaries interacted with. Clinician-level tool use in a given period was defined as at least one recorded interaction with the Chart Insights UAL activity during that period, restricted to office visits and telemedicine encounters, with the fixed cohort of 142 randomized intervention-arm clinicians as the denominator.

S3.2 Instrument cutoffs. For the Professional Fulfillment Index, overall burnout combines the work exhaustion and interpersonal disengagement domains (score ≥1.33 indicates burnout); a professional fulfillment score ≥3.0 indicates fulfillment. For the Net Promoter Score, respondents are classified as promoters (9–10), passives (7–8), or detractors (0–6).

S3.3 Modified total chart time per encounter. Chart review time was measured using a custom UAL-derived metric rather than Epic's Signal "Time in Clinical Review" metric or the tier-1 "Clinical Review" UAL activity alone. In an initial development phase, two clinicians (KL and TN) performed non-documentation-based pre-charting activities across 10 unique patients (5 patients each) while timing themselves; comparison with the Signal metric and the tier-1 UAL activity showed that neither adequately captured pre-charting time. The Signal metric was also judged insufficient because its underlying calculation was scheduled to change partway through the trial, which would have confounded observed changes over time.

In a second phase, two clinicians (AC and NZ) performed a structured pre-charting protocol for 25 additional unique patients, timing each subsection of activity to the second across chart review navigator activities, other review-oriented navigator sections (e.g., results review), and interaction with the tool itself. UAL records were then reviewed to identify the combination of activities that best approximated total observed chart review time (excluding note writing). The final metric summed all "Clinical Review" tier-1 activities (which includes "Chart Insights"), the pre-charting activity from tier-1 "Documentation," and navigator activities from tier-1 "Other". These activities captured any time before patient check out.

S3.4 Free-text feedback and categorization. Free-text feedback was collected from two sources: an open-response post-study survey item asking clinicians to describe any tool-generated inaccuracies or biases ("Please share any examples of tool-generated inaccuracies or biases you observed, including whether specific issues occurred consistently"), and unsolicited feedback submitted through the tool's embedded feedback mechanism during the intervention period. Each in-tool submission corresponded to a single generated summary so clinicians could submit multiple times within the same and across encounters. Responses from both sources were assigned to a single mutually exclusive category from an inductively developed scheme: tool limitations, inaccurate information, generality and relevance, trust barriers, omissions, and workflow and speed. For the survey item, uninterpretable or off-prompt responses were excluded, and categories were tabulated among clinicians reporting at least one concern (Table 3). In-tool submissions were categorized identically and reported descriptively (Table S1).

**Section S4. Statistical Analysis**

S4.1 Sample size justification. The sample size of 284 participants provides 80% power to detect intervention effects as small as 0.33 standard deviations (a small-to-medium effect size), assuming a two-sample t-test on pre-post change and a two-sided 0.05 significance level.

S4.2 Model specification. Linear mixed models estimated intervention effects, with random clinician effects accounting for repeated measures (baseline and follow-up for survey outcomes; baseline, period 1, and combined periods 2 and 3 for modified total chart time). Models included fixed effects for study arm, response period, and their interaction, and adjusted for clinician sex, age, specialty, number of half-days in clinic, baseline PTL, baseline modified total chart time per encounter, and ambient scribe access. Intervention effects were estimated by linear contrasts between arms at follow-up (survey outcomes) and in combined periods 2 and 3 (modified total chart time).

**Figure Legend**

**Figure S1. Contexts in which the summarization tool was reported most useful**. Intervention-arm clinicians selected up to two contexts in which they perceived the tool as most useful on the post-intervention survey; respondents could also make no selection. Values shown are the number of selections for each context.

**Table S1. Categorization of In-Tool Feedback Submissions from Intervention-Arm Clinicians**

| **Category** | **No. of submissions** | **%** |
| --- | --- | --- |
| Inaccurate information | 30 | 41.7 |
| Tool limitations | 20 | 27.8 |
| Omissions | 15 | 20.8 |
| Generality and relevance | 7 | 9.7 |
| **Total** | **72** |  |

*Unsolicited free-text feedback submitted by intervention-arm clinicians through the tool's embedded feedback mechanism during the intervention period, at the encounter level (clinicians could submit on multiple encounters). Each submission was assigned to one investigator-defined category; these categories were not native to the feedback mechanism. The coding scheme is identical to that used for Table 3 (tool limitations, inaccurate information, generality and relevance, trust barriers, omissions, workflow and speed); categories not appearing in the table produced no submissions.*
